# 3-hour genome sequencing and targeted analysis to rapidly assess genetic risk

**DOI:** 10.1101/2022.09.09.22279746

**Authors:** Miranda Galey, Paxton Reed, Tara Wenger, Erika Beckman, Irene J. Chang, Cate R. Paschal, Jillian G. Buchan, Christina M. Lockwood, Mihai Puia-Dumitrescu, Daniel R. Garalde, Joseph Guillory, Androo J. Markham, Andrew B. Stergachis, Michael J. Bamshad, Evan E. Eichler, Danny E. Miller

**Affiliations:** Division of Genetic Medicine, Department of Pediatrics, University of Washington and Seattle Children’s Hospital, Seattle, WA, USA; Department of Laboratory Medicine and Pathology, University of Washington, Seattle, WA, USA; Department of Laboratories, Seattle Children’s Hospital, Seattle, WA, USA; Department of Genome Sciences, University of Washington School of Medicine, Seattle, WA, USA; Brotman Baty Institute for Precision Medicine, University of Washington, Seattle, WA, USA; Division of Neonatology, Department of Pediatrics, University of Washington and Seattle Children’s Hospital, Seattle, WA, USA; Oxford Nanopore Technologies, Oxford, UK; Division of Medical Genetics, Department of Medicine, University of Washington, Seattle, WA; Howard Hughes Medical Institute, University of Washington, Seattle, WA, USA

## Abstract

Rapid genetic testing in the critical care setting enables targeted evaluations, directs therapies, and helps families and care providers make informed decisions about goals of care. We tested whether we could perform ultra-rapid assessment of genetic risk for a Mendelian condition, based on information from an affected sibling, in a newborn via whole-genome sequencing using the Oxford Nanopore platform. By optimization of the DNA extraction and library preparation steps paired with targeted analysis, we were able to demonstrate within three hours of birth that the newborn was neither affected nor a carrier for variants underlying acrodermatitis enteropathica. This proof-of-concept experiment demonstrates how prior knowledge of familial variants can be used to rapidly evaluate an individual at-risk for a genetic disease.

## MAIN TEXT

The benefits of rapid genetic testing in critically ill individuals have been demonstrated.^1–6^ A precise genetic diagnosis helps guide management and testing options while giving families and providers valuable information to make informed decisions about goals of care.^6–8^ Because management decisions for critically ill individuals often must be made in hours or days, minimizing the time required to make a precise genetic diagnosis is of broad interest.

The turnaround time for rapid genetic testing via whole-genome sequencing has decreased from approximately 26 hours in 2015 to just under 8 hours in early 2022.^1,4,9–11^ Reductions in the turnaround time have been enabled by advances in sequencing chemistry and the development of analysis pipelines that can quickly and efficiently prioritize variants with limited manual input. Further improvements using short-read sequencing-by-synthesis approaches are constrained by the amount of time required to perform each step of the synthesis reaction, thus new approaches are needed. Recently, Oxford Nanopore sequencing was used to rapidly evaluate a cohort of critically ill individuals with the shortest time to identification of a pathogenic variant in just under 8 hours.^1^ Nanopore technology is an ideal platform on which to develop ultra-rapid sequencing approaches because sequencing data from individual DNA molecules are available in near real-time.^12^

We report the whole-genome sequencing followed by targeted analysis of a newborn known to be at risk of a genetic condition within three hours of birth **(Figure 1)**. We set forth to further decrease the time required to identify pathogenic variants via genome sequencing by optimizing the DNA extraction and library preparation steps followed by targeted analysis based on prior genetic information. We also sought to use smaller blood volumes—an important consideration in neonates where there is an upper limit on the amount blood that may be drawn per day.^13^ Finally, because clinical adoption of ultra-rapid sequencing on the Nanopore platform will likely require automation of some steps, such as library preparation, we sought to test a protocol that required fewer manual touchpoints and therefore should be easier to automate.

**Figure 1:**
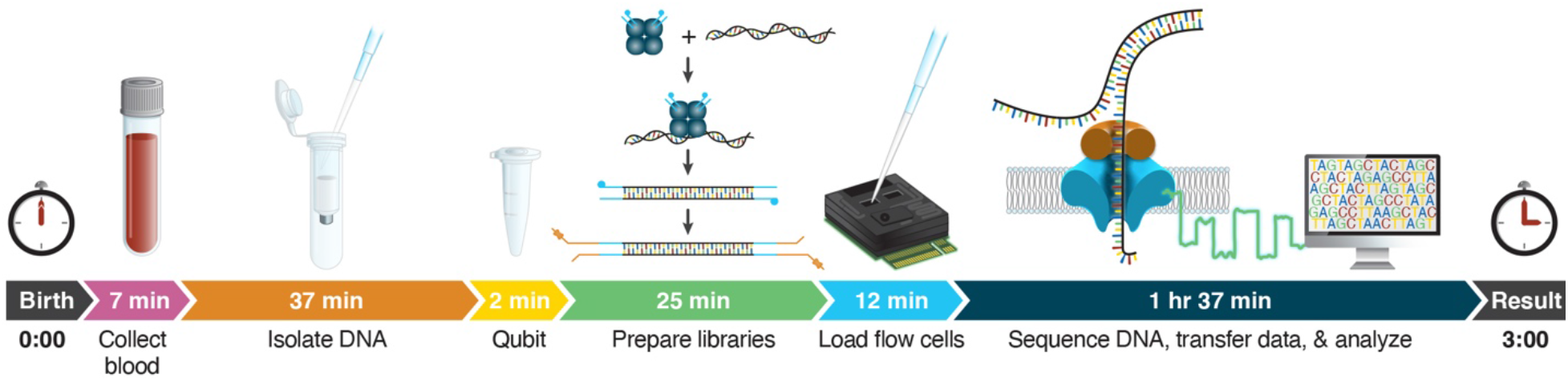
Timeline from birth to result. The proband in this report was born at hour 0. Cord blood was collected in the delivery room and walked to the laboratory. DNA isolation and QC required 39 total minutes. Library preparation and loading of 10 flow cells was completed in 37 minutes. Once the first 10 flow cells were confirmed to be running well 10 additional flow cells were loaded. Approximately 3 hours after the proband was born, Clair3 was run and reads mapping to the *SLC39A4* region were isolated and phased into two distinct haplotypes. This demonstrated that the newborn did not inherit either the known pathogenic or the candidate promoter variants.

The newborn and his genetic sibling (#1) were conceived using donated anonymized embryos. Sibling #1 developed symptoms suggesting acrodermatitis enteropathica (MIM: 201100), an autosomal recessive condition due to zinc transporter defect encoded by *SLC39A4*, that results in reduced intestinal absorption of zinc. After beginning elemental zinc supplementation sibling #1 had near complete resolution of symptoms. Sibling #1 has a persistent delay in developmental milestones.

Clinical testing of sibling #1 identified only a single pathogenic variant (NM_130849.4:c.1203G>A, p.Trp401*) in *SLC39A4*. To identify a second pathogenic variant putatively missed by clinical testing, we performed targeted long-read sequencing of blood-derived DNA from sibling #1, and found a promoter variant (c.-169A>G) on the other allele in an evolutionarily conserved CCAAT box that demonstrates chromatin accessibility and transcription factor occupancy selectively within gastrointestinal cells **(Figure 2A, S1, methods)**.^12,14–16^ The c.-169A>G variant disrupts the A at the 4th position of the CCAAT box, and is predicted to be disruptive based on several predictive algorithms (i.e. FINSURF score of 0.8936, and LINSIGHT score of 0.927754).^17,18^ CCAAT boxes are essential components of many promoters, and play a critical role in promoting transcription.^19^ Thus, it is suspected that this variant represents a “second hit” in this individual through loss of transcription factor binding to the CCAAT box, and subsequent loss of *SLC39A4* transcription from this allele. No other family members were available for testing.

**Figure 2:**
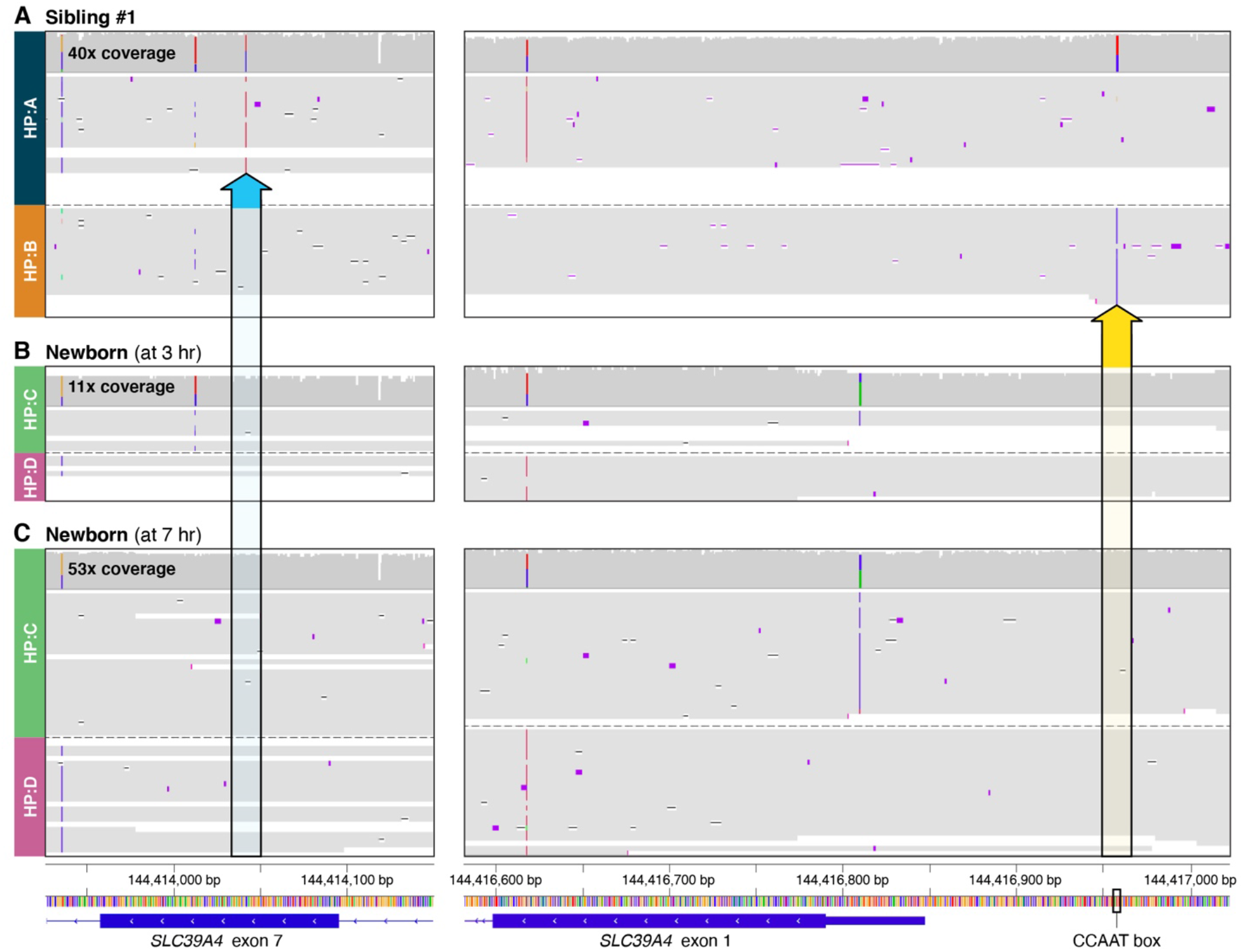
IGV view of *SLC39A4* variants found in the affected sibling (sibling #1) (A), and the newborn at 3 hours (B) and 7 hours of life (C). Haplotypes denoted on the left are as listed in Table S4. Indels of < 3 nt have been hidden. **A**. The known pathogenic c.1203G>A, p.Trp401* variant is present on HP:A (blue arrow) while the c.-169A>G promoter variant is found on HP:B (yellow arrow). **B**. Phased reads from the newborn after approximately 1 and 1/2 hours of sequencing, or at 3 hours of life. Neither the previously known pathogenic single nucleotide variant (blue box) or the promoter variant (yellow box) were observed in the newborn. **C**. After 5 and 1/2 hours of sequencing, or at 7 hours of life, neither of the previously known variants were identified. In both sibling #1 (A) and the newborn (B, C) nearly all reads span the 3 kb distance between the two variants.

Before the pathogenic genetic variants in sibling #1 were known the family elected to proceed with implantation of a randomly selected embryo from the same IVF cycle as sibling #1. Because of the uncertain impact of zinc deficiency on early development of sibling #1, and the inability to rely on biochemical testing for diagnosis during the neonatal period the family agreed to pursue rapid genetic testing after birth to determine if the newborn had inherited either or both the c.1203G>A and c.-169A>G promoter variants in *SLC39A4* found in sibling #1. The neonate was born at term by Cesarean section and the neonatal course was unremarkable. He was discharged home at 2 days of age.

Cord blood was collected at birth, and ultra-rapid whole-genome long-read sequencing was performed using 20 Nanopore PromethION flow cells with sequencing libraries prepared by a combination of two commercially available Nanopore kits **(methods)**. The advantage of the rapid library protocol is that sequencing libraries can be generated in as little as 10 minutes using a transposase to insert adapter attachment sites followed by chemical attachment of a sequencing adapter. The downside of this approach is that the random integration of the transposase into DNA can result in shorter overall DNA fragments than the ligation-based library preparation protocol. Accordingly, the average read lengths for our 20 libraries ranged from 6–11 kb with batch effects from our combined library preparation reactions **(Table S2)**.

Analysis of phased data at 3 hours post-birth (1.5 hours of sequencing) indicated that the neonate had inherited neither the pathogenic c.1203G>A, (p.Trp401*) variant nor the candidate c.-169A>G promoter variant found in sibling #1 **(Figure 2B)**, suggesting that he was not at high risk for inherited acrodermatitis enteropathica **(Table S4)**. We allowed sequencing to continue for an additional 4 hours (5.5 hours of sequencing total), generating approximately 45x coverage by 7 hours of life **(Table S6)**. Subsequent variant calling with Clair3 and phasing with LongPhase confirmed the findings from 3 hours of life **(Figure 2C)**. Approximately 83% of heterozygous variants on chromosomes 1–22 identified after 1.5 hours of sequencing were present at 5.5 hours of sequencing with 99.5% of those having been assigned to the correct haplotype **(Table S7)**.

The time required to make a precise genetic diagnosis or to evaluate an individual at risk of a known genetic condition segregating in their family could likely be further reduced by simplifying DNA extraction and library preparation steps, perhaps by automation of these steps on a single microfluidic device. Simplification of the flow cell preparation and loading steps will be required before large-scale studies of sub-4-hour genome sequencing can be carried out. Although automation of flow cell preparation and loading will be challenging, it is likely necessary for translating this approach into the clinical testing environment. Large-scale evaluation of ultra-rapid genome sequencing cannot rely on the ability of clinical laboratory staff to quickly perform the required steps and then expertly load tens of flow cells to generate sufficient data for analysis. Instead, a single device that performs DNA extraction, library preparation, and flow cell loading—as well as flow cells that produce more data per flow cell—will likely be required to make widespread ultra-rapid genome sequencing on the Nanopore platform in the clinical environment a reality.

Our analysis was simplified by several factors—a focus on a single gene, knowledge of the pathogenic and candidate variants in that gene, and knowledge of neighboring single nucleotide polymorphisms (SNP) defining the affected haplotypes in sibling #1. This is not unlike other clinical scenarios when a newborn is known to be at risk of inheriting familial variants that have caused disease in other family members. Often, sequence data is available for affected individuals and could be used in the same way we have used it here to perform rapid assessment. For those individuals in which a candidate variant or gene is unknown real-time variant calling and phasing could be performed with results compared to a database of curated variants.^1^

For some critically ill newborns with clinical findings such as hyperammonemia, severe hypoglycemia, lactic acidosis of unknown etiology, or seizures in the first hours of life, an ultra-rapid precise genetic diagnosis could be pivotal to guiding treatment decisions. For other newborns the benefit remains unclear as much of the first 6–12 hours of life involve stabilization and assessment of illness. Thus, genetic testing results may not be considered until the newborn is stabilized and the family has considered what role ultra-rapid results may have in their decision-making process.^20^ Nonetheless, the approach described here can be applied broadly to other patient populations of all ages where suspicion of a genetic etiology is high, and a precise genetic diagnosis could guide treatment choices.

## SUPPLEMENTAL FIGURES

**Figure S1:**
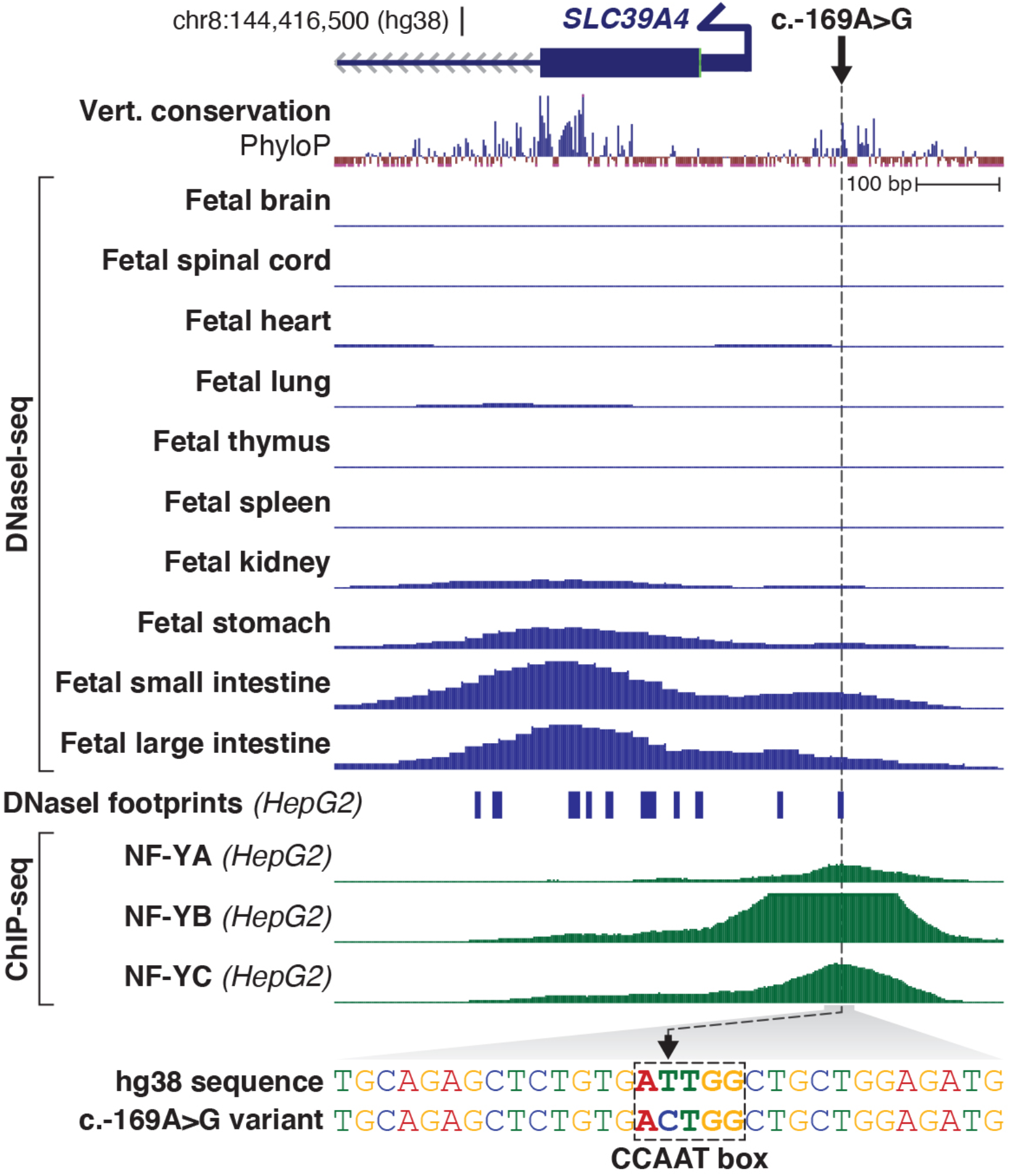
The c.-169A>G variant is located within a region that is evolutionarily conserved and demonstrates chromatin accessibility and transcription factor occupancy selectively within gastrointestinal cells.

## METHODS

### Targeted long-read sequencing and analysis of sibling #1

Sibling #1 is the sibling of the newborn and is affected with acrodermatitis enteropathica. DNA for sequencing was extracted from blood using the Monarch HMW DNA Extraction Kit for Cells & Blood (NEB #T3050) following the suggested protocol for DNA isolation from blood with the following specifications: 500 µl of blood was used as input, shaking occurred at 900 RPM, bead binding time was extended to 8 minutes, and a sterile glass plating bead was used to homogenize the elution. DNA was sheared to an average fragment length of 10 kb using a Covaris gTUBE as described previously.^16^ Libraries for sequencing were prepared using the Oxford Nanopore Ligation Kit (SQK-LSK110) and loaded onto a R9.4.1 flow cell on a GridION. MinKNOW version 21.10.8 running Guppy 5.0.17 was configured to run Adaptive Sampling with a target region of approximately 2.6 Mb for approximately 48 hr **(Table S1)**. FASTQ files were generated with Guppy 5.0.12 using the superior model (dna_r9.4.1_450bps_sup.cfg). Single nucleotide and indel variants were called with Clair3,^21^ phased with LongPhase,^22^ and annotated with VEP 103.1^23^ using annotations from SpliceAI^24^ and CADD v1.6.^25^

### DNA extraction and quantification from the newborn

At birth, 1 mL of cord blood was collected in an EDTA tube and placed on ice. DNA was extracted using the Monarch Genomic DNA Purification Kit (NEB #T3010) following the recommended protocol for genomic DNA purification from mammalian whole blood. Briefly, five individual reactions were prepared. 100 μL of whole blood was added to a 2-mL microfuge tube along with 10 μL of Proteinase K, 3 μL of RNase A, and 100 μL of Blood Lysis Buffer. This was vortexed and incubated for 5 min at 56°C in a thermal mixer at 1,400 rpm. 400 μL of gDNA binding buffer was then added to each tube and pulse-vortexed for 10 sec. The combined lysate and binding buffer were transferred to a gDNA purification column in a collection tube. The tube was centrifuged at 1,000 x g for 1 min then 12,000 x g for 3 min. Columns were transferred to a new collection tube and 500 μl of gDNA wash buffer was added, the cap was closed, and the collection tube was inverted 3 times. Tubes were then centrifuged for 1 min at 12,000 x g, the flow-through was discarded, and an additional 500 μL of gDNA wash buffer was added and centrifuged again for 1 min at 12,000 x g. The gDNA purification columns were transferred to 1.5-mL low-bind tubes and 61 μL of gDNA elution buffer preheated to 60°C was added and allowed to sit for 1 min at room temperature. The columns were then centrifuged for 1 min at 12,000 x g. 1 μL of each extraction was quantified with the Qubit dsDNA HS (High Sensitivity) Assay Kit (ThermoFisher) **(Table S2)**.

### Library preparation

Libraries for sequencing were prepared using a combination of reagents from the Oxford Nanopore Rapid Sequencing (SQK-RAD004) and PCR-cDNA Sequencing (SQK-PCS111) kits. Four separate library preparation reactions were prepared in 1.5-mL Eppendorf tubes by combining 80 μL of genomic DNA with a target of 2–4 µg of DNA per reaction **(Table S3)**. 30 μl of nuclease-free water was added to each tube to bring the total volume to 110 μL. 12.5 μL of FRA was added to each tube and incubated at 30°C for 2 min followed by 80°C for 2 min, then placed in a cold block for 15 sec. 5 µL of RAP-T from the SQK-PCS111 kit was added to each tube and incubated for 5 min at room temperature. The RAP-T sequencing adapter from the SQK-PCS111 kit was used instead of the RAP adapter from the SQK-RAD004 kit to take advantage of its higher pore occupancy rate, which results in higher output. After 5 min, 72.5 μL of nuclease-free water was added to each tube and the tube was placed on ice. The library mix for loading was made by adding 32 μL of library to a 1.5-mL Eppendorf lo-bind tube followed by 75 μL of SQB and 43 μL of nuclease-free water, for a total volume of 150 μL.

### Flow cell preparation, loading, and sequencing

During the DNA extraction and library preparation steps, 20 Oxford Nanopore R9.4.1 PromethION flow cells were prepared for sequencing. Flow cells were allowed to sit at room temperature for at least 10 min before being placed into a PromethION 24 running MinKNOW control software v22.03.4. Ten flow cells were primed with 500 μl of FB + FLT, then allowed to sit for at least 5 min before an additional 500 μl of FB + FLT was added to each. 150 μl of the library mix was then loaded to each flow cell and allowed to sit for 5 min prior to beginning sequencing. After confirming that the first 10 flow cells were working as expected a second group of 10 flow cells were primed and loaded as described. Each experiment was configured to run using the RAD004 protocol, with reserved pores and live base calling turned off.

### Data transfer and base calling

Original sequencing data was transferred to a remote base calling server with rsync. Basecalling was done using Guppy 6.2.1 (Oxford Nanopore) and the dna_r9.4.1_450bps_hac_prom.cfg model with a minimum quality score cutoff of 7 ‘--min_qscore 7’. A custom script was used to monitor for incoming sequencing data and perform sequential basecalling of each library on a single NVIDIA A100 GPU. Basecalling was performed on a machine with four NVIDIA A100 GPUs, thus four libraries could be called simultaneously. After the first round of basecalling for each library, the ‘--resume’ flag was used to resume basecalling from the last position for each library.

### Alignment and analysis

A custom script monitored each library for new FASTQ files generated by the basecalling process. New FASTQ files were combined into a single FASTQ file then aligned to GRCh38 using minimap2.^26^ After alignment, SAMtools was used to extract reads surrounding the target gene, *SLC39A4* (chr8:14,400,000-14,450,000) and two control genes; *COL1A1* (chr17:50,000,000-50,300,000) and *F8* (chrX:154,800,000-155,200,000).^27^ Individual BAM files for each target gene were merged with previous BAM files using SAMtools merge. Beginning at 3 hr after birth and every 30 min thereafter, variants were called on the combined bam file with Clair3^21^ followed by phasing using LongPhase.^22^ The phased bam file was then visualized using IGV.^28^

The newborn was evaluated for inheritance of the known pathogenic variants beginning approximately 1 hr after sequencing started, or 2.5 hr after birth. Visual inspection at that time did not reveal either the pathogenic or candidate promoter variant. At 3 hr after birth variants called by Clair3 in the interval chr8:144,411,754-144,419,728 were compared to those found in the affected sibling **(Table S4)** and suggested that the newborn did not carry either the pathogenic C>T at chr8:144,414,042 or the candidate promoter T>C at chr8:144,416,958 **(Figure 2B)**. Sequencing was stopped 7 hours after birth, or 5 and 1/2 hours after sequencing started **(Table S5)**, and all flow cells were washed and stored. Single nucleotide variants (SNV) identified at 3 hours of life and their haplotype assignment remained unchanged at 7 hours of life, with the exception of a single SNV present on HP:D that was not called at 3 hours of life (chr8:144,413,427). Average coverage of chromosome 8 at 1-hour timepoints was monitored during sequencing **(Table S6)**. Variant calling statistics, phasing statistics, and switch errors were calculated using WhatsHap **(Table S7)**.^29^

### Study approval and consent

The study was approved by the University of Washington institutional review board and consent was obtained for each participant. The legal representatives of both individuals described in this report consented to having the results of this research work published.

## Data Availability

Data that support the findings of this study are available upon request from the corresponding author. Additional clinical information about the individuals described here are available upon request from the corresponding author.

## SUPPLEMENTAL TABLES

**Table S1:** Targets used for adaptive sampling of the sibling (#1) affected with acrodermatitis enteropathica. Coordinates are for GRCh38.

| Gene Target | Chromosome | Target start | Target end | Target size (bp) |
| --- | --- | --- | --- | --- |
| *SLC39A4* | chr8 | 143,400,000 | 145,450,000 | 2,050,000 |
| *COL1A1* | chr17 | 50,150,000 | 50,250,000 | 100,000 |
| *FMR1* | chrX | 147,850,000 | 148,000,000 | 150,000 |
| *AR* | chrX | 67,500,000 | 67,800,000 | 300,000 |

**Table S2:** Concentrations after DNA extraction and quantification, including which library preparation reaction the DNA was used in.

| gDNA extraction | Quantification (ng/μl) | Total DNA (μg) | Library added to (volume added) |
| --- | --- | --- | --- |
| Tube 1 | 49.4 | 4.0 | Library 1 (60 μl) |
| Tube 2 | 49.4 | 4.0 | Libraries 1 (20 μl) & 2 (40 μl) |
| Tube 3 | 25.4 | 2.0 | Libraries 2 (40 μl) & 3 (40 μl) |
| Tube 4 | 57.0 | 4.6 | Libraries 3 (40 μl) & 4 (20 μl) |
| Tube 5 | 52.8 | 4.2 | Library 4 (60 μl) |

**Table S3:** Total starting DNA concentrations for each library, the group of flow cells that the library was loaded on, and output statistics for each library group.

| Library | Total DNA input (μg) | Flow cells library loaded on | Ave read length (kb) | Average bases per library group (Gb) |
| --- | --- | --- | --- | --- |
| Library 1 | 4.0 | 1–5 | 6.3 | 8.5 |
| Library 2 | 3.0 | 6–10 | 6.9 | 10.0 |
| Library 3 | 2.2 | 11–15 | 8.6 | 9.3 |
| Library 4 | 4.3 | 16–20 | 10.3 | 9.0 |

**Table S4:**
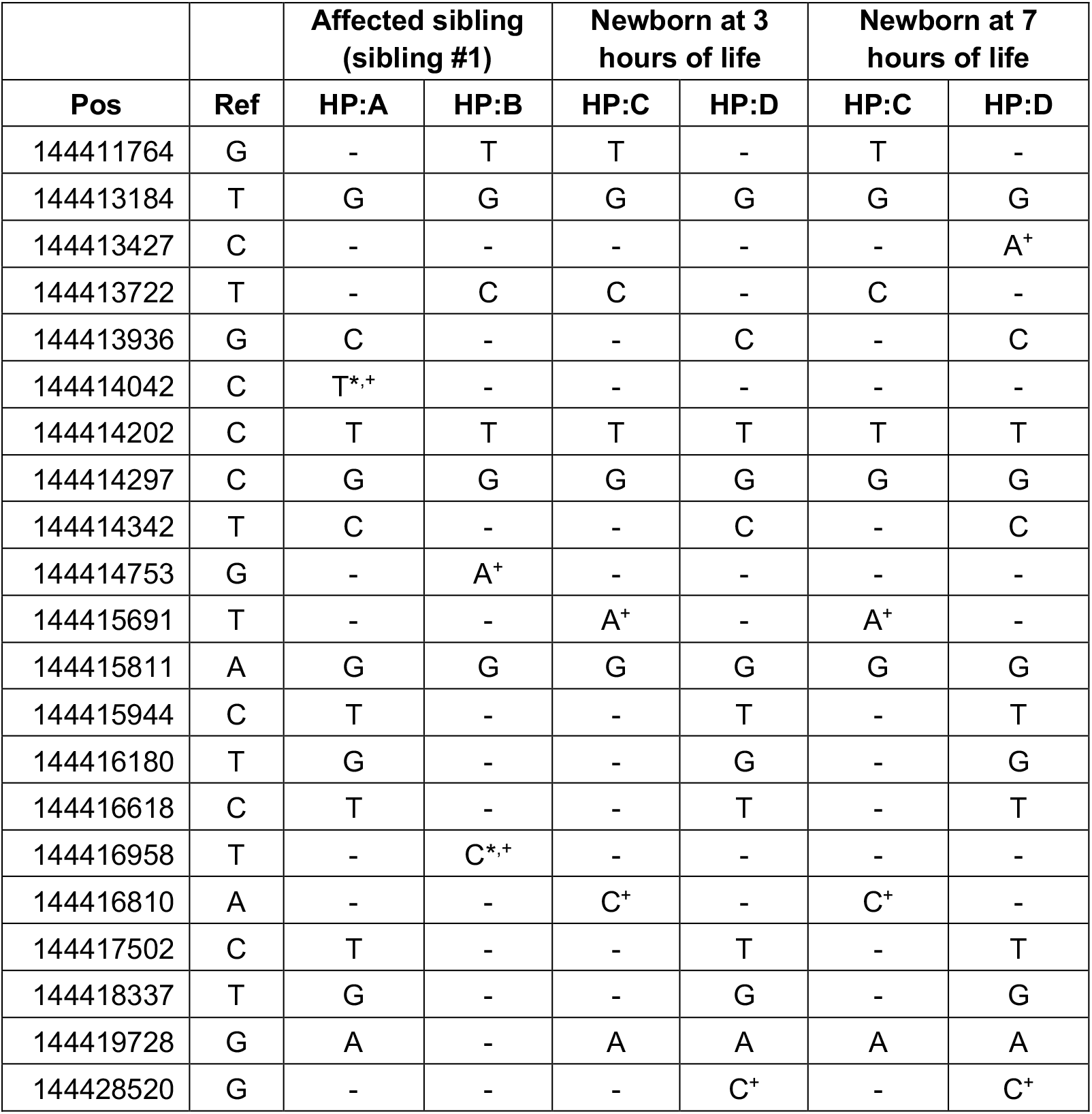
SNPs present on chromosome 8 within an 8-kb region that includes *SLC39A4*, as well as a more distant SNP at 144,428,520, which supports differentiation of HP:A and HP:D. Asterisks denote the pathogenic and candidate variant identified in the affected sibling (#1), plus signs denote polymorphisms unique to the given haplotype. HP, haplotype.

**Table S5:** Per-flow cell sequencing statistics. Library preparation was divided into four groups. Flow cells 1–10 were loaded first and once data quality was validated flow cells 11–20 were loaded.

| Flow cell | Library group | Total bases (Gb) | Average read length (kb) | Reads (millions) | Runtime (h:mm) |
| --- | --- | --- | --- | --- | --- |
| 01 | Library 1 | 8.7 | 5.9 | 3.4 | 3:52 |
| 02 | Library 1 | 8.8 | 6.1 | 3.2 | 3:49 |
| 03 | Library 1 | 8.1 | 6.1 | 3.0 | 3:48 |
| 04 | Library 1 | 9.3 | 6.9 | 3.2 | 3:47 |
| 05 | Library 1 | 7.5 | 6.5 | 2.7 | 3:46 |
| 06 | Library 2 | 9.3 | 6.8 | 3.0 | 3:46 |
| 07 | Library 2 | 9.9 | 7.0 | 3.2 | 3:51 |
| 08 | Library 2 | 11.1 | 6.9 | 3.5 | 3:49 |
| 09 | Library 2 | 10.4 | 6.9 | 3.4 | 3:48 |
| 10 | Library 2 | 9.2 | 6.7 | 3.1 | 3:47 |
| 11 | Library 3 | 9.7 | 8.5 | 2.7 | 3:22 |
| 12 | Library 3 | 9.2 | 8.6 | 2.6 | 3:20 |
| 13 | Library 3 | 8.4 | 8.4 | 2.5 | 3:20 |
| 14 | Library 3 | 9.7 | 8.4 | 2.8 | 3:25 |
| 15 | Library 3 | 9.4 | 9.0 | 2.5 | 3:23 |
| 16 | Library 4 | 9.1 | 10.8 | 2.0 | 3:22 |
| 17 | Library 4 | 9.3 | 10.0 | 2.3 | 3:20 |
| 18 | Library 4 | 8.6 | 10.4 | 2.0 | 3:17 |
| 19 | Library 4 | 8.8 | 10.3 | 2.1 | 3:16 |
| 20 | Library 4 | 9.1 | 10.1 | 2.2 | 3:16 |

**Table S6:**
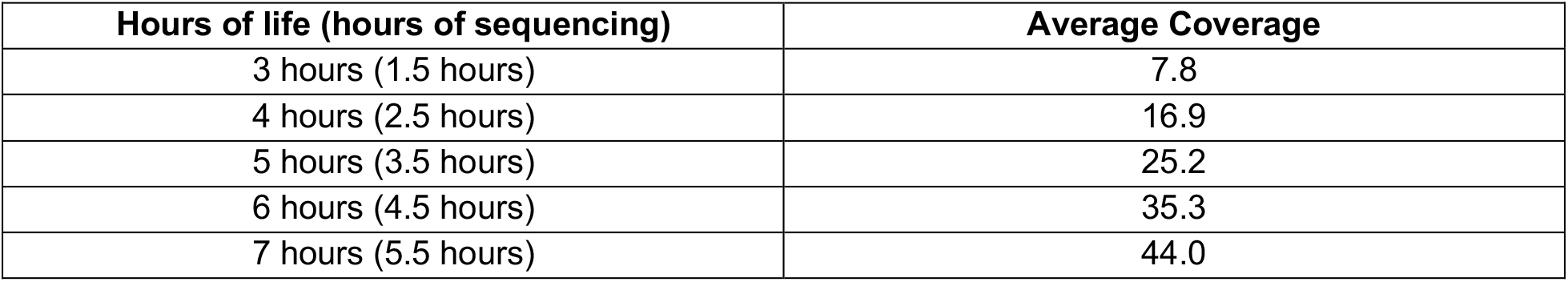
Average coverage of chromosome 8 at 1-hour timepoints.

| Hours of life (hours of sequencing) | Average Coverage |
| --- | --- |
| 3 hours (1.5 hours) | 7.8 |
| 4 hours (2.5 hours) | 16.9 |
| 5 hours (3.5 hours) | 25.2 |
| 6 hours (4.5 hours) | 35.3 |
| 7 hours (5.5 hours) | 44.0 |

**Table S7:** SNVs called and phased at 3 hours of life (1.5 hours of sequencing) compared to those at 7 hours of life (5.5 hours of sequencing). Percent of variants at 3 hours of life was calculated by dividing the shared heterozygous variants by the total heterozygous variants identified at 3 hours of life. Phasing switches are the number of variants that switched from one haplotype assignment at 3 hours of life to the other haplotype at 7 hours of life. Chromosomes X and Y were excluded from this analysis.

| Chr | Heterozygous variants at 3 hours of life | Heterozygous variants at 7 hours of life | Shared heterozygous variants | Percent of variants at 3 hours | Phasing switches | Switch error rate |
| --- | --- | --- | --- | --- | --- | --- |
| chr1 | 169923 | 190747 | 142846 | 84% | 688 | 0.5% |
| chr2 | 187993 | 207690 | 158457 | 84% | 614 | 0.4% |
| chr3 | 152144 | 169843 | 129026 | 85% | 435 | 0.3% |
| chr4 | 160875 | 193764 | 137174 | 85% | 733 | 0.5% |
| chr5 | 137973 | 158521 | 117864 | 85% | 486 | 0.4% |
| chr6 | 145206 | 158656 | 124055 | 85% | 292 | 0.2% |
| chr7 | 133887 | 148902 | 111488 | 83% | 431 | 0.4% |
| chr8 | 117096 | 135676 | 100895 | 86% | 375 | 0.4% |
| chr9 | 107231 | 129170 | 87152 | 81% | 635 | 0.7% |
| chr10 | 120866 | 132025 | 100745 | 83% | 611 | 0.6% |
| chr11 | 105658 | 123933 | 90812 | 86% | 243 | 0.3% |
| chr12 | 102509 | 120163 | 85988 | 84% | 256 | 0.3% |
| chr13 | 79204 | 86226 | 68555 | 87% | 144 | 0.2% |
| chr14 | 71248 | 79287 | 60602 | 85% | 327 | 0.5% |
| chr15 | 67383 | 85113 | 56416 | 84% | 392 | 0.7% |
| chr16 | 75881 | 83682 | 61439 | 81% | 380 | 0.6% |
| chr17 | 68352 | 73110 | 53443 | 78% | 299 | 0.6% |
| chr18 | 64512 | 82473 | 55478 | 86% | 182 | 0.3% |
| chr19 | 53035 | 57284 | 42244 | 80% | 247 | 0.6% |
| chr20 | 66058 | 70955 | 49799 | 75% | 265 | 0.5% |
| chr21 | 39956 | 42489 | 30355 | 76% | 337 | 1.1% |
| chr22 | 43293 | 46216 | 31653 | 73% | 432 | 1.4% |

## ACKNOWLEDGEMENTS

We thank the family for participating in this study and Angela Miller for editorial and figure preparation assistance. This article is subject to HHMI’s Open Access to Publications policy. HHMI lab heads have previously granted a nonexclusive CC BY 4.0 license to the public and a sublicensable license to HHMI in their research articles. Pursuant to those licenses, the author-accepted manuscript of this article can be made freely available under a CC BY 4.0 license immediately upon publication. This work was supported, in part, by a trainee grant to DEM from the Brotman Baty Institute for Precision medicine, and a US National Institute of Mental Health (NIMH) grant R01MH101221 to EEE. EEE is an investigator of the Howard Hughes Medical Institute.

